# Early prognostication of overall survival for pediatric diffuse midline gliomas using MRI radiomics and machine learning: a two-center study

**DOI:** 10.1101/2023.11.01.23297935

**Authors:** Xinyang Liu, Zhifan Jiang, Holger R. Roth, Syed Muhammad Anwar, Erin R. Bonner, Aria Mahtabfar, Roger J. Packer, Anahita Fathi Kazerooni, Miriam Bornhorst, Marius George Linguraru

## Abstract

**Background:** Diffuse midline gliomas (DMG) are aggressive pediatric brain tumors that are diagnosed and monitored through MRI. We developed an automatic pipeline to segment subregions of DMG and select radiomic features that predict patient overall survival (OS).

**Methods:** We acquired diagnostic and post-radiation therapy (RT) multisequence MRI (T1, T1ce, T2, T2 FLAIR) and manual segmentations from two centers of 53 (internal cohort) and 16 (external cohort) DMG patients. We pretrained a deep learning model on a public adult brain tumor dataset, and finetuned it to automatically segment tumor core (TC) and whole tumor (WT) volumes. PyRadiomics and sequential feature selection were used for feature extraction and selection based on the segmented volumes. Two machine learning models were trained on our internal cohort to predict patient 1-year survival from diagnosis. One model used only diagnostic tumor features and the other used both diagnostic and post-RT features.

**Results:** For segmentation, Dice score (mean [median]±SD) was 0.91 (0.94)±0.12 and 0.74 (0.83)±0.32 for TC, and 0.88 (0.91)±0.07 and 0.86 (0.89)±0.06 for WT for internal and external cohorts, respectively. For OS prediction, accuracy was 77% and 81% at time of diagnosis, and 85% and 78% post-RT for internal and external cohorts, respectively. Homogeneous WT intensity in baseline T2 FLAIR and larger post-RT TC/WT volume ratio indicate shorter OS.

**Conclusions:** Machine learning analysis of MRI radiomics has potential to accurately and non-invasively predict which pediatric patients with DMG will survive less than one year from the time of diagnosis to provide patient stratification and guide therapy.

**KEY POINTS:**

- Automatic machine learning approach accurately predicts DMG survival from MRI
- Homogeneous whole tumor intensity in baseline T2 FLAIR indicates worse prognosis
- Larger post-RT tumor core/whole tumor volume ratio indicates worse prognosis

**IMPORTANCE OF STUDY:** Studies of pediatric DMG prognostication have relied on manual tumor segmentation from MRI, which is impractical and variable in busy clinics. We present an automatic imaging tool based on machine learning to segment subregions of DMG and select radiomic features that predict overall survival. We trained and evaluated our tool on multisequence, two-center MRIs acquired at the time of diagnosis and post-radiation therapy. Our methods achieved 77-85% accuracy for DMG survival prediction. The data-driven study identified that homogeneous whole tumor intensity in baseline T2 FLAIR and larger post-therapy tumor core/whole tumor volume ratio indicates worse prognosis. Our tool can increase the utility of MRI for predicting clinical outcome, stratifying patients into risk-groups for improved therapeutic management, monitoring therapeutic response with greater accuracy, and creating opportunities to adapt treatment. This automated tool has potential to be easily incorporated in multi-institutional clinical trials to provide consistent and repeatable tumor evaluation.

## Introduction

Diffuse midline gliomas (DMG), including diffuse intrinsic pontine gliomas (DIPG), are aggressive central nervous system pediatric tumors located in the brainstem and thalamus.^1^ As one of the most devastating pediatric cancers, DMG represents about 10–15% of all pediatric tumors of the central nervous system, with an estimated 300 new cases diagnosed annually in the USA.^2^ Most DMGs occur between the ages of 5 and 10 years, with a peak at 7 years.^3^ There is no curative therapy for DMG, and radiation therapy (RT) is the standard treatment with only transitory benefits.^4^ Despite numerous clinical trials of new agents and novel therapeutic approaches over the last decades,^5^ disease outcomes remain dismal with a median overall survival (OS) of less than 1 year, a 2-year OS rate of less than 10%,^6^ and a 5-year OS rate of less than 1%.^7^

Magnetic resonance imaging (MRI) is the standard noninvasive test for DMG diagnosis and monitoring of tumor response to therapy. Although pediatric DMGs have a diverse imaging appearance,^8^ MRI features have been used to predict H3K27M mutation status^9^ and correlate with patient prognosis.^10-15^ The features utilized in these studies were either low-dimensional image features^10,11,13-15^ or based on texture analysis.^12^ The statistical analyses that most of these studies relied on tend to identify inconsistent and inconclusive imaging biomarkers across different studies and datasets. For example, a study of 357 pediatric DIPGs demonstrated that although many MRI features, such as tumor size, enhancement and necrosis etc., were strongly associated with survival on univariable analysis, very few were significantly associated with survival on multivariable analysis.^11^ These findings suggest that only relying on statistical analysis of conventional MRI findings may not be sufficient to predict OS in DMGs.

Machine learning has shown great potential to predict survival or discriminate between certain groups in studies of other brain tumors such as glioblastoma multiforme (GBM) and pediatric low-grade gliomas.^16-19^ For DMG, machine learning-based regression models were proposed to correlate with patient prognosis based on extracted MRI radiomic features.^20,21^ These studies only focused on imaging data at diagnosis, and the tumors were segmented manually, which is generally believed to be time-consuming and to have high inter-operator variability. Other studies demonstrated that semiautomated DMG volume measurements are more accurate, prognostically relevant, and consistent than manual measurements.^14,15^ In addition to diagnostic scans, it is also important to consider longitudinal data at post-treatment timepoints.^10^

With new therapeutic strategies currently under investigation for DMG, including epigenetic therapy and immunotherapy,^22^ there is a great need for noninvasive prognostic imaging tools that can be universally used to accurately identify which patients are at risk for the most rapid deterioration, and thereby assist clinical trial eligibility and therapy planning. Such tools should be automatic, objective, and easy to use in multi-institutional clinical trials. With the vast advancements in deep learning techniques, there has been tremendous success in automatic segmentation of brain tumors from MRI, including adult,^23,24^ pediatric brain tumors,^25,26^ and our previous work of segmenting DMG^27,28^. These advancements have the potential to enable us to create a fully automatic, image-based radiomic analysis and DMG prognostic tool.

In this work, we developed a novel imaging tool to process and analyze DMG patient’s MRI data with the goal of predicting their 1-year OS. One year is the median OS of our internal cohort, and it is also close to the median OS of 11 months reported on larger DIPG studies.^11^ Therefore, accurate prediction of patient’s 1-year OS could have profound impact on the clinical management of DMG. The proposed tool is automatic, and it provides deep learning-based segmentation of subregions of DMG from MRI, radiomic feature extraction and selection based on the segmented volumes, and machine learning-based OS prediction. The proposed method was trained and validated on an internal cohort from Children’s National Hospital (CNH) to investigate the accuracy of OS prediction in 1) a baseline study using MRIs obtained at diagnosis, and 2) a post-RT study using MRIs obtained at both diagnosis and post-RT (i.e., after the first RT). The method was further tested on an external DMG dataset from Children’s Hospital of Philadelphia (CHOP) to assess the reproducibility of our findings.

## Materials and Methods

### Study Cohort

For this two-center retrospective study, institutional review board approval was obtained at both participating institutions (CNH IRB Protocols #1339 and #14310; the Children’s Brain Tumor Network (CBTN), ^29^ IRB requirement waived). Our internal cohort from CNH includes 53 pediatric and adolescent patients diagnosed with DMG between 2005-2022 (F=29, M=24) at CNH. The median patient age at diagnosis is 6.5 years with a range of 3.2–25.9 years. The median OS is 12 months with a range of 3.3–132 months from diagnosis (1 patient is still alive).

The external cohort from CHOP includes 16 pediatric patients diagnosed with DMG between 2005-2022 (F=9, M=7), made available by CBTN. The median age at diagnosis is 9.4 years with a range of 3.8–18.2 years. The median OS is 9.6 months with a range of 1.3–27.1 months from diagnosis.

### MRI Data

Both institutions used scanners and imaging protocols that varied among patients and timepoints because of retrospective data collection. For each patient, 4 MRI sequences at diagnosis and/or post-RT were collected including T1-weighted (T1), contrast-enhanced T1 (T1ce), T2-weighted (T2), and T2-weighted-Fluid-Attenuated Inversion Recovery (T2 FLAIR). The MRIs were acquired either on 1.5T or 3T magnet, with 2D or 3D acquisition protocols, using scanners from GE Healthcare, Siemens AG, or Toshiba. T1 and T1ce MRIs included T1 SE, T1 FSE, T1 MPRAGE, or T1 SPGR. T2 MRI included T2 SE, T2 FSE, T2 FRFSE or T2 propeller. T2 FLAIR MRI included those with or without gadolinium (Gd) enhancement. The slice thickness range was 0.5–6 mm and matrix range was (256–512)×(256–512) pixels. All images were collected in the DICOM image format.

Manual segmentation of DMG volumes was used as the ground truth for training the deep learning segmentation model. It was performed under the supervision of two expert neurooncologists using ITK-SNAP.^30^ Inter-expert variability was resolved through consensus. Because necrosis/cyst is not consistently identifiable for DMG, two labels were created: tumor core (TC) and whole tumor (WT). TC included two components: the Gd-enhancing tumor appearing as enhancement on T1ce, and the necrotic/cystic core appearing as hypointense on T1ce. WT includes TC and the peritumoral edematous/infiltrated tissue appearing as abnormal hyperintense signal on T2 FLAIR.

### Automatic DMG Segmentation

Despite the tremendous success of deep learning-based automatic segmentation for adult GBMs, the direct application of these methods on rare pediatric brain tumors remains challenging^31^. While GBMs and DMGs share several clinical properties, they have distinctive characteristics as well, especially in their location in the brain and radiologic presentation. Our approach was to transfer knowledge learnt from GBM segmentation to DMG segmentation.

The Brain Tumor Segmentation (BraTS) challenge is an ongoing annual event that has been held since 2012. We obtained imaging data of 1,251 GBM patients that was publicly available from BraTS.^32^ For each patient, 4 MRI sequences (T1, T1ce, T2, and T2 FLAIR) and manual segmentations of TC and WT subregions of GBM were provided. The winning method of the BraTS 2020 challenge was based on nnU-Net^24^, a popular and robust semantic deep-learning segmentation method. nnU-Net analyzes the training data and automatically configures a matching U-Net^33^-based segmentation pipeline.

Figure 1 shows the model architecture of our transfer learning-based approach using nnU-Net. It includes a pretraining phase of nnU-Net using the GBM dataset. Because nnU-Net automatically determines the segmentation pipeline based on the specific dataset, we changed this pretraining paradigm to first design the segmentation pipeline based on the DMG dataset, and then used the planned pipeline to perform pretraining on the GBM data. The pretrained network weights were then used as initialization to finetune the model using the DMG dataset. Preprocessing was performed in an automatic fashion and included N4 bias correction to correct for MRI inhomogeneities^34^, rigid registration to the SRI-24 Atlas for spatial alignment^35^, and skull stripping^36^. The output of the segmentation model was the TC and WT volumes, which were used as input to the radiomic feature extraction step.

**Figure 1.**
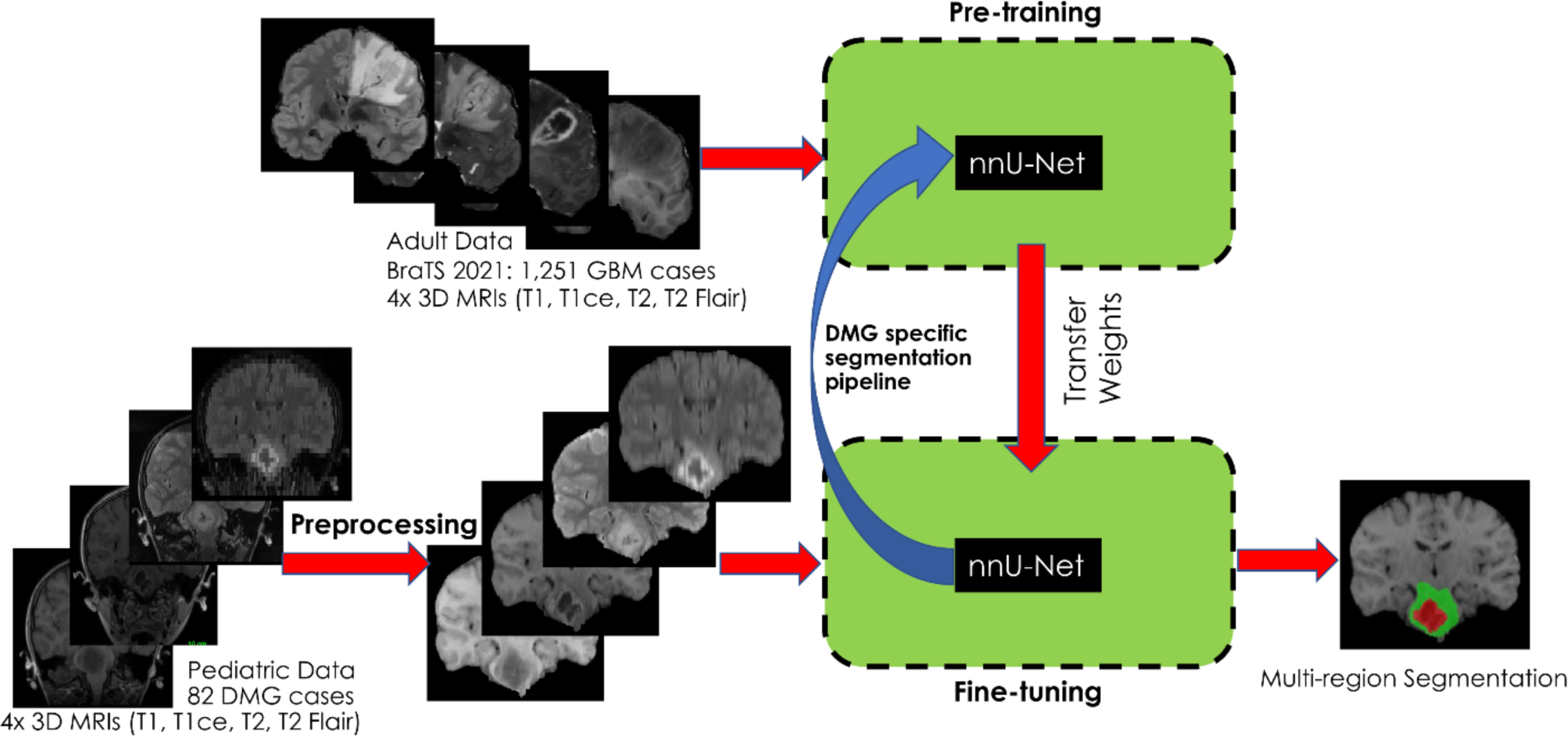
Model architecture of our DMG segmentation method, which employs nnUnet-based pre-training and fine-tuning. The input to pre-training is the adult brain tumor dataset from the BraTS 2021 challenge. The input to fine-tuning is the preprocessed DMG dataset. Based on the DMG dataset, a specific segmentation pipeline is determined and used for pre-training. After pre-training, the obtained weights are used as input for fine-tuning. The output of the model is multi-region segmentation masks.

### Experiments and Evaluation for Tumor Segmentation

Data from 45 CNH patients (with manual segmentation) were used for training and validation of the segmentation model. Scans at diagnosis and post-RT of the same patient were counted as separate MRI sets for the purpose of segmentation, each set containing four MRI sequences. This yielded a total of 82 sets from the 45 patients. Specifically, 41/82 sets were acquired at diagnosis, 34/82 sets were acquired within 1-month post-RT, and the rest of 7 sets were acquired 2–4 months post-RT.

The 82 DMG sets were randomly divided into 5 folds, and 5-fold cross-validation was performed to obtain the TC and WT volumes. Data from the same patient was always kept in the same fold. Dice coefficient and volume similarity were used as evaluation metrics to compare the predicted and ground truth segmentations, where the volume similarity is calculated as the ratio between the smaller of the compared volumes and the average of the compared volumes^37^. After 5-fold cross-validation, we trained a final model with all 82 sets and used it to predict TC and WT volumes for the remaining 8 internal patients and 16 external patients.

Many DMG cases do not have or have very small TC volumes. Thus, comparison between predicted and ground truth in small or absent TC volumes produces extreme metrics (e.g., Dice score of 0 or 1). To void bias to small volumes, we cleaned predicted volumes by removing small (i.e., <130 mm^3^) disconnected regions. Moreover, let TC/WT denote the ratio between TC volume and WT volume. We did not evaluate segmentation performance if 0<TC/WT<4% for both predicted and ground truth segmentations. If TC/WT=0 for both, the metrics were set to be 1. The thresholds of 130 voxels and 4% were determined by a previous study on the pediatric brain tumor data.^38,39^

### Radiomic Feature Extraction

Based on automatically segmented DMG volumes, we used the open-source PyRadiomics software^40^ to extract radiomic features including 13 volumetric and shape features and 91 gray level features. Please refer to Supplemental Appendix S1 for a complete list of features. The gray level features included: 18 first order features, 22 gray level co-occurrence matrix (GLCM) features, 16 gray level size zone matrix (GLSZM) features, 16 gray level run length matrix (GLRLM) features, 5 neighboring gray tone difference matrix (NGTDM) features, and 14 gray level dependence matrix (GLDM) features. In addition, we added two demographic features (i.e., sex and age), and two volumetric features of interest (i.e., brain volume and relative tumor volume [DMG volume divided by brain volume]). Because gray level features are susceptible to inter-scanner variation due to different acquisition protocol^41^, image gray levels were normalized by removing the mean and scaling to unit variance before the features were calculated.

The baseline study employed 401 features, including sex, age, 35 volumetric and shape features, and 4 sets of 91 gray level features (one set for each MRI sequence). The volumetric and shape features include brain volume, 14 WT features (i.e., 13 from PyRadiomics and relative DMG volume), 10 TC features, and 10 features for the ratio between TC and WT (TC/WT). Because many DMG cases do not have TC volume, four features (i.e., elongation, flatness, surface area to volume ratio, and sphericity) having measurements of TC in the denominator of their calculation were excluded. The gray level features were calculated based on WT segmentations.

The post-RT study employed 1,576 features, including sex, age, 118 volumetric and shape features and 1,456 gray level features. The volumetric and shape features include brain volumes at diagnosis and post-RT, 28 WT features (14 at diagnosis and 14 post-RT), changes of 14 WT features (post-RT values minus values at diagnosis), relative changes of 14 WT features (changes divided by values at diagnosis), 20 TC features (10 at diagnosis and 10 post-RT), changes of 10 TC features, 20 TC/WT features (10 at diagnosis and 10 post-RT), and changes of 10 TC/WT features. We did not include relative changes of TC and TC/WT features because measurements related to TC at diagnosis could be null, which would make the definition of relative change invalid. The gray level features included 4 sets of 91 gray level features at diagnosis, 4 sets of 91 gray level features post-RT, changes of 4 sets of 91 gray level features, and relative changes of 4 sets of 91 gray level features.

### Feature Selection

Feature selection was performed on the training data prior to prediction to avoid overfitting. In the first step, feature filtering was performed using the Mann-Whitney U test comparing feature values between short OS (<1 year) and long OS (≥1 year). Sixty-nine features with p<0.05 were selected for the post-RT study. For the baseline study, because there was only 1 feature with p<0.05, we selected 10% of all features (40 features) with the smallest p-values. Sequential feature selection was then performed on the filtered features to select the optimal number of discriminative features for each study. Let *n* be the desired number of features, which we capped at 10% of the number of patients to avoid overfitting the model to the training data. The algorithm added 1 feature at an iteration to form a feature subset in a greedy fashion until *n* was reached. At each iteration, the algorithm went through each feature not currently in the feature subset and chose the feature to add such that the new feature subset achieved the best accuracy in the leave-one-out cross-validation. Specifically, we trained a linear support vector machine (SVM) to classify between short OS and long OS using all subjects in our internal cohort except for 1, which was used for testing. This process was repeated iteratively until all patients were tested. Because of our small datasets, we used leave-one-out cross-validation to maximize the number of training examples, and employed the linear kernel for SVM, which is less prone to overfitting than non-linear kernels.

### Experiments and Evaluation for OS Prediction

Images at diagnosis of 52/53 CNH patients were used for training and validation in the baseline study. There were 26/52 patients with short OS, i.e., survival shorter than one year from diagnosis. One patient did not have images of all 4 MRI sequences at diagnosis, but the post-RT images were used for training the segmentation model. Images at diagnosis and within 3 months post-RT of 41/52 patients were available and used for training and validation in the post-RT study. There were 22/41 patients with short OS.

After feature selection, the final SVM model was trained with all internal patients with the selected features. Validation of the final model on the internal dataset was reported. The final model was used to predict OS based on the same selected features on the external dataset. For the baseline study, 16 external patients (9 had short OS) were tested. 9/16 external patients who had post-RT imaging (<3 months) were tested in the post-RT study. 4/9 patients had short OS.

## Results

### Segmentation Results

Table 1 shows performance of the automatic DMG segmentation method evaluated on the internal and external datasets. The external evaluation shows performance on out-of-distribution data to reflect generalizability based on scanning and protocol variability and tumor heterogeneity^42^. Metrics of WT segmentation for the external cohort (0.86 mean Dice score and 0.91 mean volume similarity) were similar to those obtained for the internal cohort (0.88 mean Dice score [Mann-Whitney U test p=0.10] and 0.93 mean volume similarity [p=0.13]). This suggests our method can be successfully generalized for segmenting WT volume of images from different sources. Similarly, metrics of TC segmentation for the external cohort (0.74 mean Dice score and 0.81 mean volume similarity) were similar, although inferior to those obtained for the internal cohort (0.91 mean Dice score [p=0.10] and 0.93 mean volume similarity [p=0.58]). Note that the median Dice score (0.83) and volume similarity (0.99) of TC segmentation for the external cohort were improved from the mean and the model performed well on TC segmentation for most external cases (12/14).

**Table 1.** Mean (median) and standard deviation of Dice coefficient and volume similarity calculated by comparing predicted tumor core (TC) and whole tumor (WT) volumes and those segmented manually. Results shown include validation on the internal cohort (from Children’s National Hospital) and testing on the external cohort (from Children’s Hospital of Philadelphia).

| Evaluation dataset | TC Dice | WT Dice | TC vol. similarity | WT vol. similarity |
| --- | --- | --- | --- | --- |
| Internal cohort | 0.91 (0.94) ± 0.12 | 0.88 (0.91) ± 0.07 | 0.94 (0.99) ± 0.10 | 0.93 (0.96) ± 0.07 |
| External cohort | 0.74 (0.83) ± 0.32 | 0.86 (0.89) ± 0.06 | 0.81 (0.99) ± 0.34 | 0.91 (0.93) ± 0.07 |

Figure 2 shows qualitative segmentation results on the diagnosis and post-RT images of a DMG patient of the internal cohort. The Dice scores for this case were 0.92 (diagnostic TC), 0.92 (diagnostic WT), 0.97 (post-RT TC), and 0.93 (post-RT WT), which were approximately the median Dice scores for our internal cohort (0.94 for TC and 0.91 for WT in Table 1).

**Figure 2.**
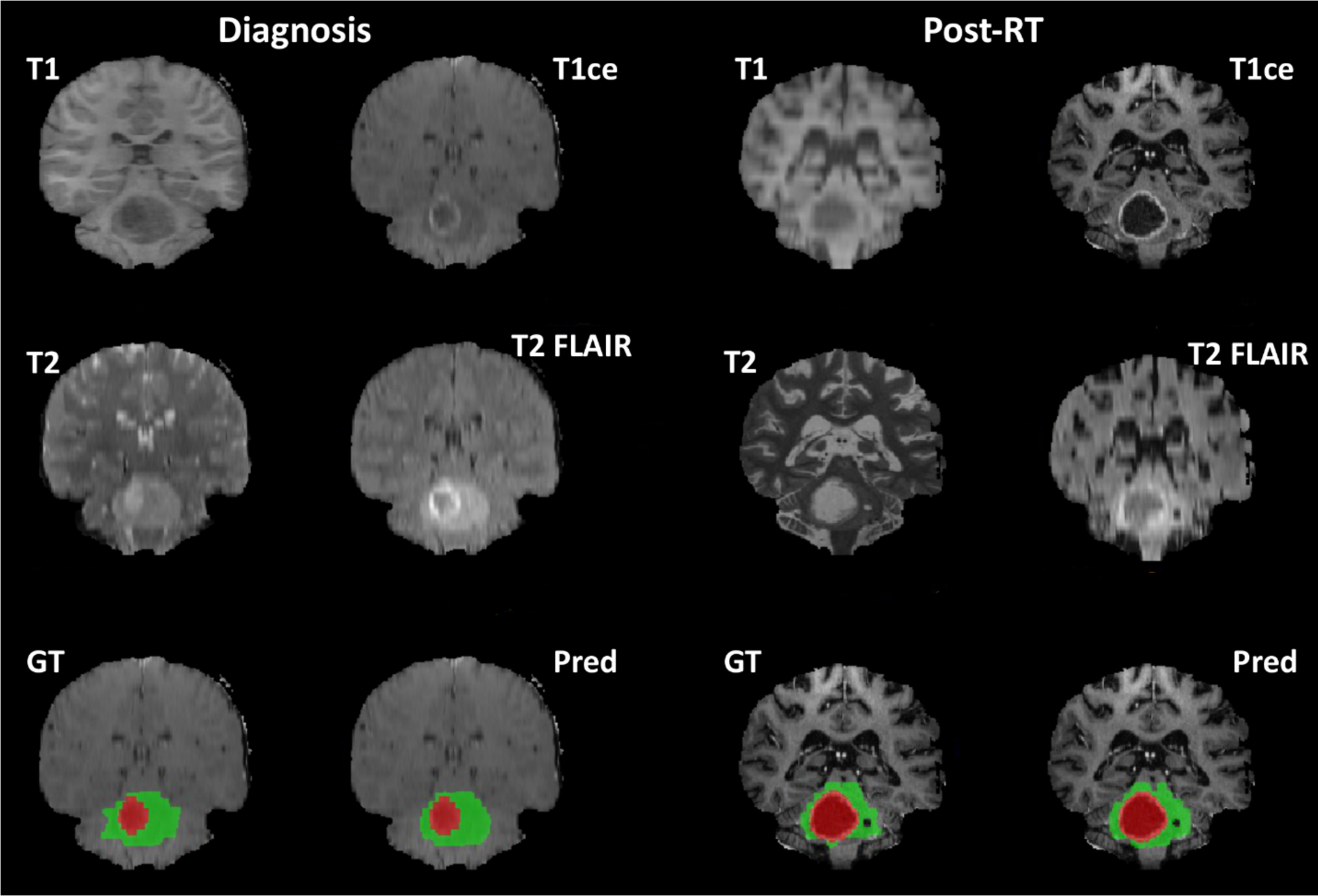
Qualitative segmentation results on the diagnosis and post-RT images of a DMG patient from the internal cohort. The figure shows 4 MRI sequences after preprocessing, the ground truth (GT) segmentation, and the predicted (Pred) segmentation generated by our method (red: tumor core volume, red + green: whole tumor volume).

### OS Prediction Results

Table 2 shows results of the proposed OS prediction method. Because identifying patients with higher risk (i.e., OS<1 year) is critical, we adjusted model parameters to maximize accuracy or sensitivity. In general, the results suggest that adding post-RT data may improve prediction accuracy and sensitivity over the baseline. Despite the small data cohort, the evaluation metrics on our external cohort were generally comparable to those obtained on the internal cohort, indicating overall generalizability of our machine learning predictive model.

**Table 2.** Results of the proposed OS prediction method. Short OS of less than one year is considered positive. We present results at the operating points of maximum accuracy and maximum sensitivity.

| Study | Accuracy | Sensitivity | Specificity | Accuracy | Sensitivity | Specificity |
| --- | --- | --- | --- | --- | --- | --- |
|  | Internal cohort (52 subjects) |  |  | External cohort (16 subjects) |  |  |
| **Baseline max accuracy** | 77% | 81% | 73% | 81% | 89% | 71% |
| **Baseline max sensitivity** | 62% | 92% | 31% | 75% | 100% | 43% |
|  | Internal cohort (41 subjects) |  |  | External cohort (9 subjects) |  |  |
| **Post-RT max accuracy <sup>a</sup>** | 85% | 100% | 68% | 78% | 100% | 60% |

<sup>a</sup> Also max sensitivity

The number of selected features for the baseline and post-RT studies was 5 and 4, respectively. We list below the selected features for each study, along with their interpretation for prediction and the p-values of Mann-Whitney *U* test between short and long OS computed on our internal cohort. The features are listed in the order of their relevance to OS prediction.

The 5 selected features for the baseline study are:

- GLCM Information measure of correlation (Imc1) on T2 FLAIR (p=0.118): quantifies the complexity of the texture. It ranges from −1 to 0 and the higher the value the more complex in texture.
- GLSZM High gray level zone emphasis on T1 (p=0.231): measures the distribution of the higher gray level values.
- The median gray level value on T2 FLAIR (p=0.173)
- Skewness on T2 (p=0.061): measures the asymmetry of the distribution of gray level values about the mean value.
- The 10^th^ percentile of gray level value on T2 FLAIR (p=0.217)

The significant feature for the baseline study is the GLCM Cluster Shade on T2, which is a measure of skewness and uniformity (p=0.009). However, our feature selection algorithm did not select this feature. This verifies our method selects features that perform best in combination in the machine learning approach, but not necessarily the features with the smallest p-values.

The 4 selected features for the post-RT study are:

- The ratio of maximum 2D diameter (coronal plane) between post-RT TC and post-RT WT (p=0.017). The maximum 2D diameter is the largest pairwise Euclidean distance between tumor surface mesh vertices on a 2D plane.
- The 10^th^ percentile of gray level value on post-RT T1ce (p=0.027).
- The ratio of minor axis length between post-RT TC and post-RT WT (p=0.002). The minor axis length is the second-largest axis length of principal component analysis performed on the volume.
- Root mean squared on post-RT T1ce (p=0.006): is the square-root of the mean of all the squared gray level values.

Figure 3 shows the comparison between short OS and long OS for the selected features of the 2 studies. A visual example of radiomics is shown in Fig. 4.

**Figure 3.**
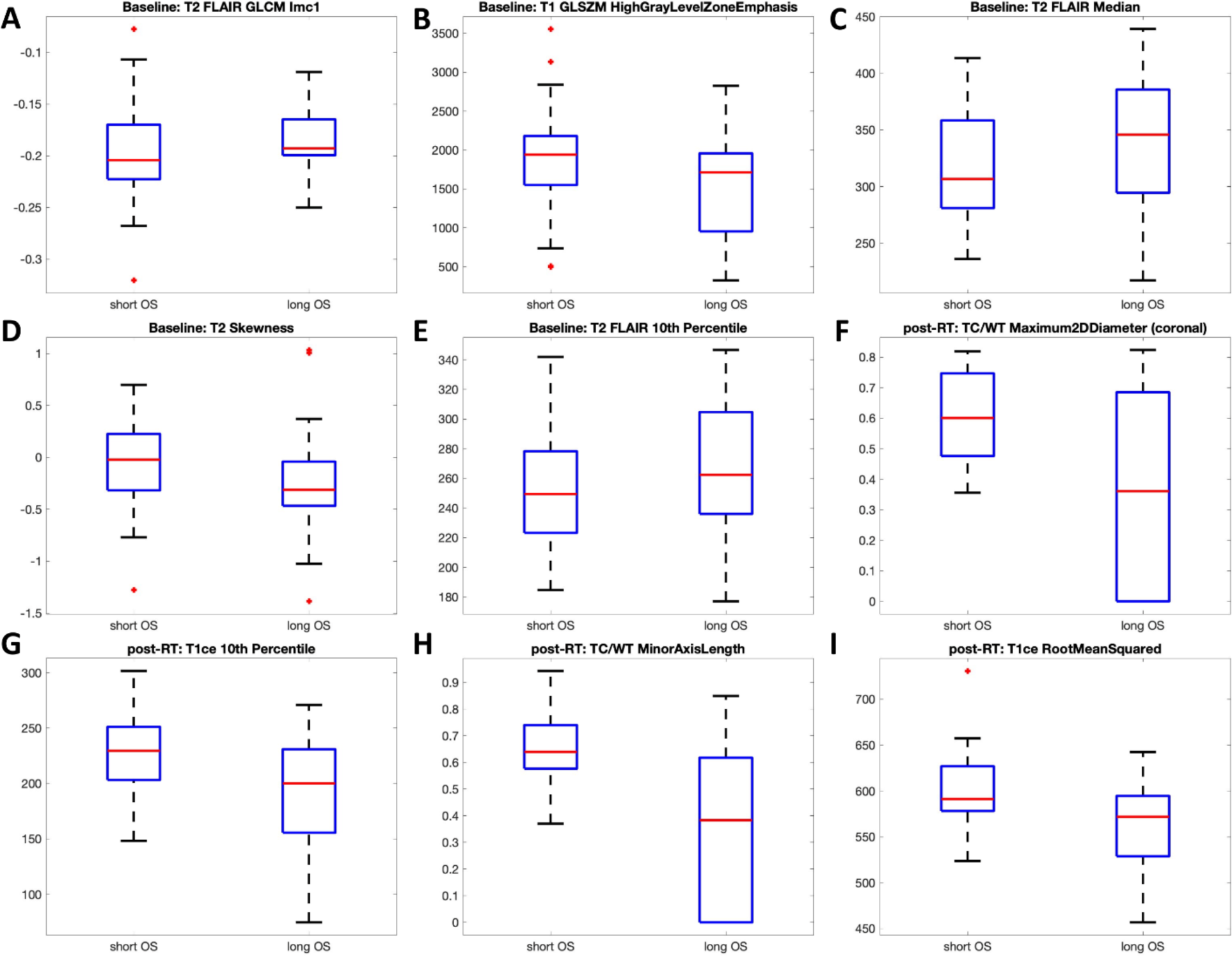
Comparison between short OS and long OS for the selected features of the baseline (A–E) and the post-RT (F–I) studies. Data of both internal and external cohorts were considered. **A**: GLCM Imc1on T2 FLAIR; **B**: GLSZM high gray level zone emphasis on T1; **C**: median gray level on T2 FLAIR; **D**: skewness on T2; **E**: 10th percentile gray level on T2 FLAIR; **F**: the ratio between TC and WT for maximum 2D diameter in the coronal plane; **G**: 10th percentile gray level on T1ce; **H**: the ratio between TC and WT for minor axis length; **I**: root mean square of gray level on T1ce.

**Figure 4.**
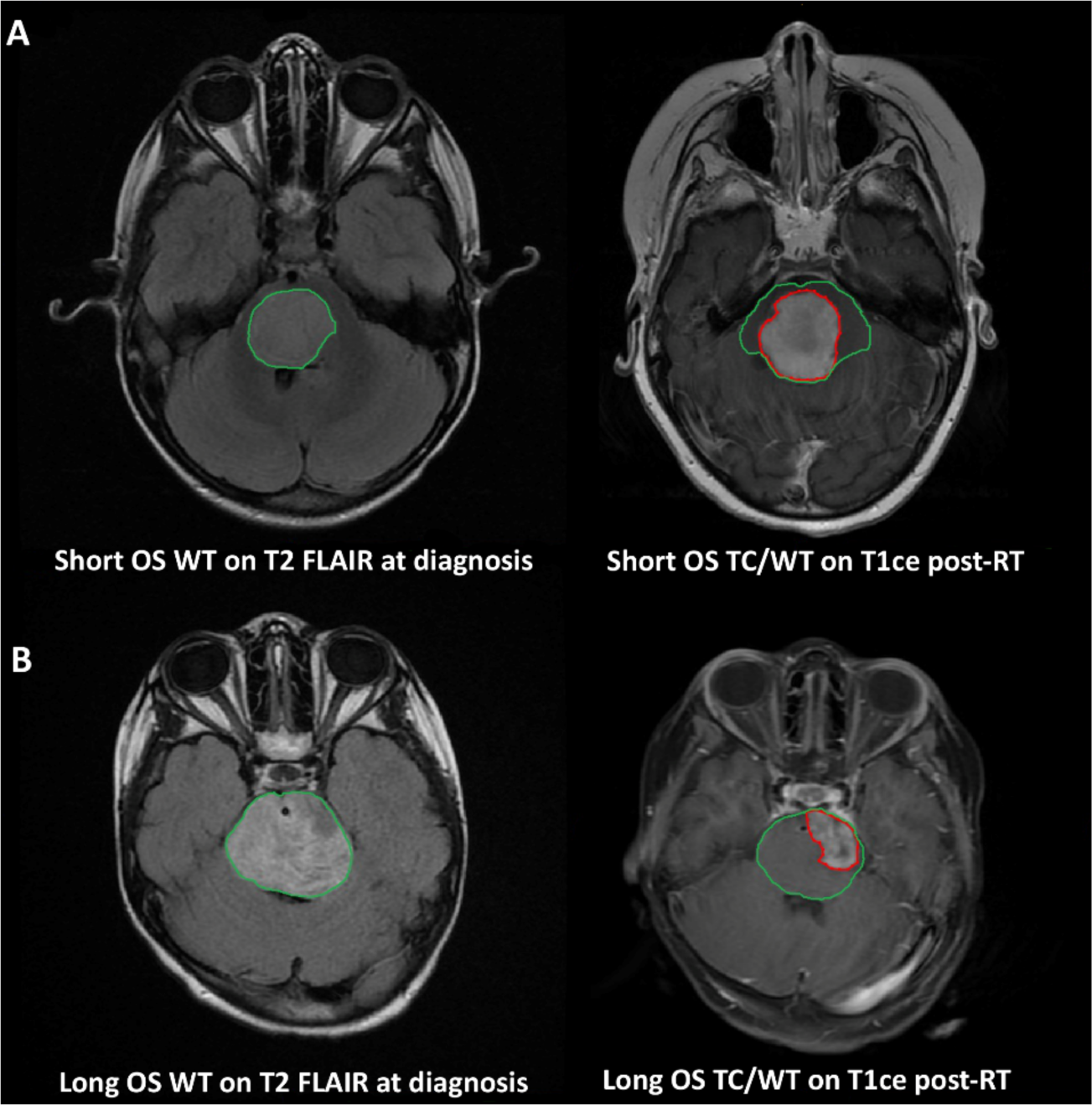
MRI of two patients who survived 8 months (A, short OS) and 14 months (B, long OS) from our internal cohort. Manual segmentations were outlined (red: tumor core [TC], green: whole tumor [WT]). Diagnostic T2 FLAIR suggests in WT intensity distribution is more homogeneous and intensity values are relatively lower (in contrast to nearby tissues) for short OS compared with long OS. Post-RT T1ce suggests the TC/WT ratio of short OS is larger than that of long OS. These observations were consistent with our findings in the selected features (Fig. 3).

## Discussion

There are no standardized machine learning-based tools avaible to clinics to provide quantitative radiomic and predictive analytics for pediatric brain cancers. The analysis of brain tumors on MRI, and especially of rare pediatric tumors, has been challenged by small data cohorts acquired by different scanners and imaging protocols, and by manual segmentations with inter-observer variability. Machine learning models have the potential to extract complex imaging patterns, provide automation and standardization for the analysis, and support the evaluation of clinical trials—and ultimately of patient therapy—with repeatable and consistent data.

To our best knowledge, this study is the first to report a fully automatic, machine learning-based model to prognosticate DMG survival using MRI features. Our automatic DMG segmentation method generated accurate TC and WT segmentations. The mean Dice scores of 0.91 for TC and 0.88 for WT obtained on the internal cohort were comparable to those reported for adult GBM segmentation using state-of-the-art deep learning models.^43,44^ The results on the external cohort were similar for WT, an indication of model generalizability and robustness when applied to independent data with different imaging and patient characteristics. Although results were inferior for TC segmentation for the external cohort (mean Dice=0.74), they are comparable to the 0.62–0.74 Dice scores reported in a recent study of automatic segmentation of subregions of pediatric brain tumors^26^. Our results are also comparable with results of the winning method (TC Dice=0.78, WT Dice=0.82)^38^ for pediatric brain tumor segmentation challenge 2023.^39^

A recent study based on manual tumor segmentation presented a machine learning-based regression model to correlate MRI radiomic features with DIPG prognosis.^20^ The study employed T1ce and T2 MRI acquired at diagnosis, and found that homogeneous tumor pixel intensity or texture, such as the GLCM features, conferred a shorter OS. A similar pattern was found in our baseline study, where tumors in the short OS group tend to have more homogeneous gray level distribution (i.e., smaller value of GLCM Imc1) as shown in Fig. 3A and Fig. 4.

Although diagnostic features were considered in the post-RT study, all the selected features in the post-RT study were related to post-RT measurements. Tumor volumetric and shape features, which are independent of scanner variation, were selected for the post-RT study, whereas no shape feature was selected for the baseline study. These results suggest post-RT features are more discriminative and potentially more robust compared with diagnostic features. The two selected shape features in the post-RT study indicate that larger post-RT TC/WT ratio predicts OS shorter than one year (Fig. 3FH). For both baseline and post-RT studies, our method predicted short OS with high sensitivity and specificity for both internal and external cohorts.

Our study is not without limitations. Both of our internal and external cohorts are small datasets, which is a challenge for studies of rare diseases. The findings of this study should be further verified with a larger DMG dataset. Given the data size, we used machine learning predictors based on SVM, which perform better on small cohorts and offer feature interpretability. Better DMG segmentation and OS prediction models can be achieved by training on a larger dataset, and the fully automatic nature of the proposed method is well suited for such large multi-institutional collaboration. Another potential limitation is the fact that radiomics are susceptible to bias and variation due to inter-scanner factors such as different acquisition protocols. We addressed this limitation by normalizing the distribution of gray level values. Additional feature harmonization methods besides what was performed in our study could be used to remove scanner effects in brain MRI radiomic features.^41,45^

In conclusion, we presented a fully automatic machine learning-based approach to compute radiomic biomarkers of DMGs from multisequence MRI. The approach can accurately and non-invasively predict overall survival for DMG patients and can be extended to other rare pediatric brain tumors. Our approach offers several advantages over the current standards of evaluation of pediatric brain tumors on MRI. Quantitative image analysis, including volumetrics of tumor components, can support the evaluation of tumor progression and response to treatment. Early prognostication of overall survival can guide patient risk stratification and clinical decisions. With automated and standardized analysis, the machine learning tool can provide data-driven evidence to clinical trials and personalized treatment strategies. These benefits combined with new histological and molecular findings could lead to progress in finding curative therapy for pediatric brain tumors.

## AUTHORSHIP

Conception and design: XL, ZJ, MB, MGL

Data acquisition: XL, ERB, AM, RJP, AFK, MB

Data analysis/interpretation: XL, ZJ, HRR, SMA, MGL

Drafting/revising critically: All

Final approval: All

## AKNOWLEDGMENTS

We would like to acknowledge Dr. Javad Nazarian, PhD, who is the PI of IRB Pro #1339 that was used to identify patients from Children’s National Hospital for this study, and to the Children’s Brain Tumor Network for making available the data from Children’s Hospital of Philadelphia. We would also like to acknowledge the patients and their families who consented to this research.

## Data Availability

All data included in the present work are not available for public use.

