## Supplemental Appendix S1 for "Early prognostication of overall survival for pediatric diffuse midline gliomas using MRI radiomics and machine learning: a two-center study"

**SUPPLEMENTAL MATERIAL**

**Appendix S1: all features used in the study**

Features from PyRadiomics
13 volumetric and shape features
- Voxel volume
- Surface area
- Surface area to volume ratio
- Sphericity
- Maximum 3D diameter
- Maximum 2D diameter (Slice)
- Maximum 2D diameter (Column)
- Maximum 2D diameter (Row)
- Major axis length
- Minor axis length
- Least axis length
- Elongation
- Flatness

*Gray level features (total 91)*
18 first order features
- Energy
- Total energy
- Entropy
- Minimum
- 10th percentile
- 90th percentile
- Maximum
- Mean
- Median
- Interquartile range
- Range
- Mean absolute deviation
- Robust mean absolute deviation
- Root mean square
- Skewness
- Kutosis
- Variance
- Uniformity

22 gray level cooccurrence matrix (GLCM) features
- Autocorrelation
- Joint average
- Cluster prominence
- Cluster shade
- Cluster tendency
- Contrast
- Correlation
- Difference average
- Difference entropy

- Difference variance
- Joint energy
- Joint entropy
- Information measure of correlation (IMC) 1
- Information measure of correlation (IMC) 2
- Inverse difference moment
- Inverse difference moment normalized
- Inverse difference
- Inverse difference normalized
- Inverse variance
- Maximum probability
- Sum entropy
- Sum of squares

16 gray level zone matrix (GLSZM) features
- Small area emphasis
- Large area emphasis
- Gray level non-uniformity
- Gray level non-uniformity normalized
- Size-zone non-uniformity
- Size-zone non-uniformity normalized
- Zone percentage
- Gray level variance
- Zone variance
- Zone entropy
- Low gray level zone emphasis
- High gray level zone emphasis
- Small area low gray level emphasis
- Small area high gray level emphasis
- Large area low gray level emphasis
- Large area high gray level emphasis

16 gray level run length matrix (GLRLM) features
- Short run emphasis
- Long run emphasis
- Gray level non-uniformity
- Gray level non-uniformity normalized
- Run length non-uniformity
- Run length non-uniformity normalized
- Run percentage
- Gray level variance
- Run variance
- Run entropy
- Low gray level run emphasis
- High gray level run emphasis
- Short run low gray level emphasis
- Short run high gray level emphasis
- Long run low gray level emphasis
- Long run high gray level emphasis

5 neighboring gray tone difference matrix (NGTDM) features

- Coarseness
- Contrast
- Busyness
- Complexity
- Strength

14 gray level dependence matrix (GLDM) features
- Small dependence emphasis
- Large dependence emphasis
- Gray level non-uniformity
- Dependence non-uniformity
- Dependence non-uniformity normalized
- Gray level variance
- Dependence variance
- Dependence entropy
- Low gray level emphasis
- High gray level emphasis
- Small dependence low gray level emphasis
- Small dependence high gray level emphasis
- Large dependence low gray level emphasis
- Large dependence high gray level emphasis

<u>Other features</u>
2 clinical features
- Sex
- Age

2 volumetric features
- Brain volume
- Relative tumor volume
